# *BDNF-DT and BDNF-AS-DT*: Novel Genes in the *BDNF* locus

**DOI:** 10.1101/2025.07.10.25331311

**Authors:** Svitlana V. Bach, Giovanna Punzi, Nuri E. Smith, Sreya Mukherjee, Joo Heon Shin, Qiang Chen, Geo Pertea, Leonardo Collado-Torres, Kristen R. Maynard, Stephanie C. Page, Joel E. Kleinman, Thomas M. Hyde, Daniel R. Weinberger, Keri Martinowich, Gianluca Ursini

**Author notes:** These authors contributed equally to this work (Svitlana V. Bach, Giovanna Punzi). Corresponding authors: Keri Martinowich, Gianluca Ursini, 855 North Wolfe Street, Suite 300, Lieber Institute for Brain Development, Johns Hopkins University Medical Campus, Baltimore, Maryland, USA.

## Abstract

Divergent transcription from bidirectional promoters is frequently observed in eukaryotic genomes, but the biological relevance of divergent RNA transcripts (DT) is unknown. We identified and characterized *BDNF-DT*, a novel DT gene, and *BDNF-AS-DT*, a novel readthrough gene, in the locus containing *BDNF*, a gene with key roles in neuronal development, differentiation, and synaptic plasticity. *BDNF-DT* is independent from the known *BDNF* antisense (*BDNF-AS*), and its expression is developmentally regulated and positively correlated with *BDNF* in human postmortem dorsolateral prefrontal cortex (DLPFC). *BDNF-DT* and *BDNF-AS-DT* expression increase after induced depolarization, but the temporal dynamics follow expression of *BDNF,* suggesting a regulatory role. Moreover, CRISPR-mediated upregulation of *BDNF* in human neural progenitor cells drives *BDNF-DT* expression. Finally, *BDNF-DT* shows higher expression in DLPFC from patients diagnosed with schizophrenia compared to neurotypical controls, and genetically predicted lower expression of the *BDNF-AS-DT* readthrough transcript is associated with schizophrenia and with the schizophrenia-associated C allele of the rs6265 single-nucleotide polymorphism. These data suggest that *BDNF-DT* and *BDNF-AS-DT* contribute to *BDNF* regulation and schizophrenia risk.

## Introduction

Divergent transcription – the phenomenon of RNA polymerase II transcribing in opposing directions from active promoters - has been described in many eukaryotic organisms (*1*). Protein-coding/long non-coding RNA (lncRNA) pairs resulting from divergent transcription of bidirectional promoters are often observed among transcriptionally regulated protein-coding transcripts (*2, 3*). Such recurring phenomena raise the possibility that divergent transcription fine-tunes regulation of protein-coding RNAs. Divergent RNAs (divRNAs) contribute to many aspects of gene expression, from transcription to translation to RNA degradation, and are involved in both activating and repressing *cis* (local) and *trans* (distant) gene expression (*1, 3–13*). In addition, divRNAs that partially overlap protein-coding genes may share features of antisense transcripts (*14*) and mediate effects on translation of their coupled protein-coding genes. DivRNAs contribute to intergenic transcription, together with readthrough transcription, a phenomenon that occurs when RNA polymerase II fails to recognize termination signals and continues transcribing beyond the gene’s end (*15–17*).

Cells of neural origin are highly transcriptionally active (*18*). DivRNAs are poised to contribute to molecular complexity in the brain because they represent a substantial amount of the lncRNAs that are expressed in transcriptionally active cells (*19*), and because they mediate diverse regulatory and functional roles in transcriptional, post-transcriptional, epigenetic, and nuclear processes (*1, 3–14*). However, while regulation by divRNAs represents a potential mechanism for tuning expression and splicing of protein-coding RNAs that are important for brain development and function, functional characterization of brain-expressed divRNAs remains limited.

Brain-derived neurotrophic factor (*BDNF*) encodes a secretory protein that plays key roles in neuronal development, differentiation and synaptic plasticity (*20*), and is implicated in multiple neuropsychiatric disorders (*21*). Conflicting results regarding BDNF associations with disease (*22*) are complicated by difficulty in disentangling the complex regulation of *BDNF* transcription. *BDNF* contains at least 9 unique promoters, which are spliced to a common 3’ coding exon, generating at least 22 transcripts encoding an identical BDNF protein. The locus also comprises several differentially spliced lncRNAs of the antisense *BDNF-AS* gene (*23*) (**Fig.1**). While BDNF is of significant interest as a therapeutic target, there are caveats associated with globally targeting BDNF signaling. For example, alterations in BDNF signaling map to discrete brain regions for different diseases (*21*). In addition, BDNF operates within a narrow window of optimal concentration to support trophic signaling in neurons. Indeed, its levels must be tightly regulated to prevent excessive neural activity, and overexpression can result in hyper-excitability and neuronal death (*24–28*).

**Fig. 1.**
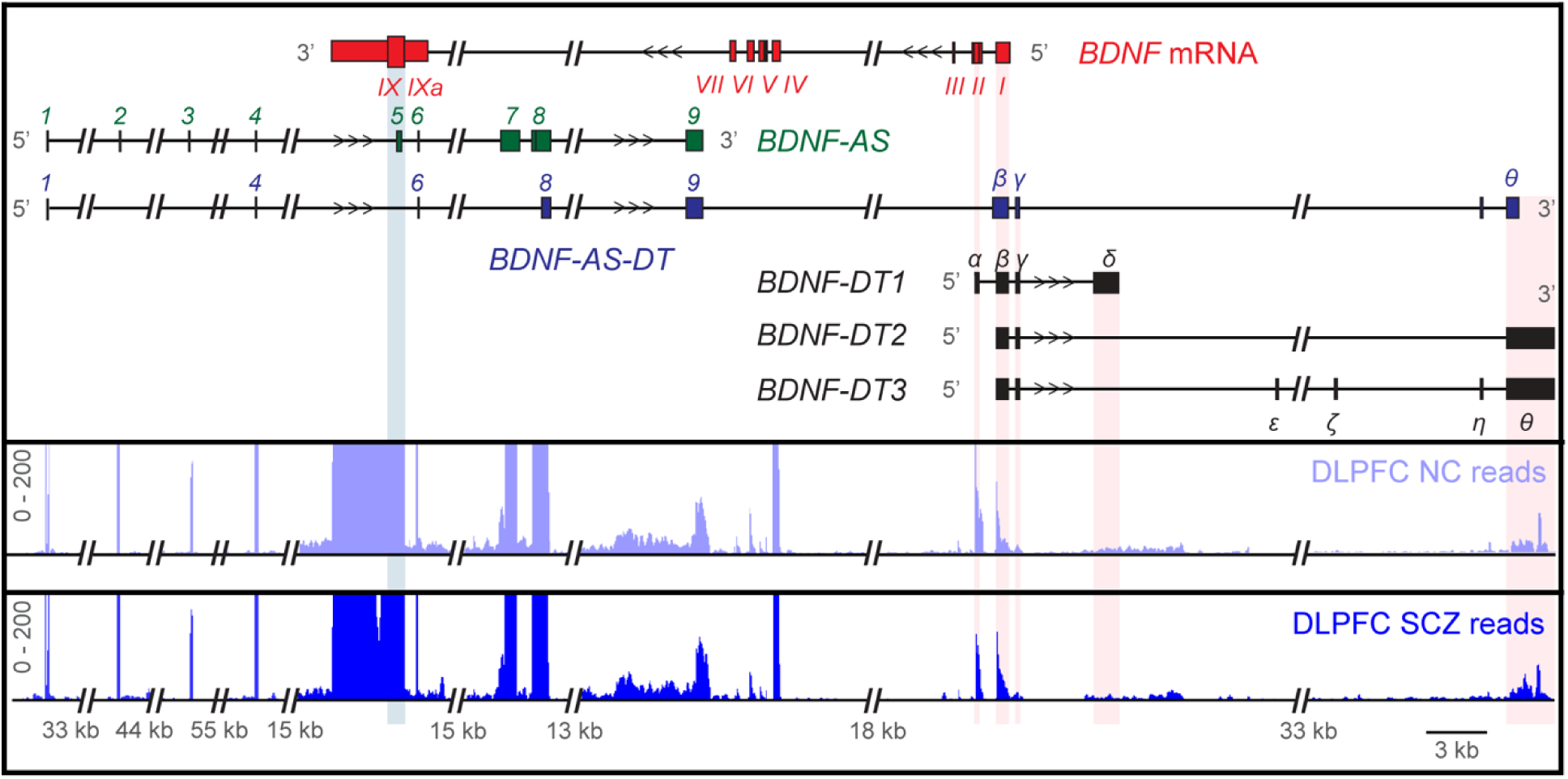
Structure, location, and expression of *BDNF-DT* and *BDNF-AS-DT* transcripts relative to *BDNF* and *BDNF-AS* in the human genome. (**Top**) *BDNF* mRNA structure with corresponding exons (*I - IXa*; red) and the coding exon *IX* (wide red box). *BDNF-AS* structure with corresponding exons (*1-9*; green). *BDNF-AS-DT* readthrough transcript structure with corresponding exons (*1*, *4*, *6*, *8*, *9*, *β*, *γ*, and *θ*; blue). *BDNF-DT1*, *2*, and *3* transcript structure with corresponding exons (*α - θ*; black). (**Bottom**) RNAseq read coverage track from bulk DLPFC postmortem human brain tissue in neurotypical controls (NC; lavender) and patients with schizophrenia (SCZ; navy), illustrating expression of *BDNF-DT* exons (pink horizontal lines) vs. *BDNF* coding exon *IX* (light blue horizontal line). Scale bar on the lower right is 3 kb.

We identified evidence for divergent transcription in the *BDNF* locus by analyzing a large RNA sequencing (RNAseq) dataset from postmortem human brain tissue (*29*) (**Fig.1**). We went on to rigorously characterize *BDNF* divergent transcripts (*BDNF-DT*) and establish activity-dependent kinetics of *BDNF* and its divergent transcripts in both human and mouse cells. We further mechanistically determined whether expression of *BDNF* from endogenous promoters alters divergent transcription. Our findings provide evidence of the existence of novel genes in the *BDNF* locus: *BDNF-DT* and the *BDNF-AS-DT* readthrough gene that is associated with genetic risk for schizophrenia. Our data suggest that *BDNF-DT* and *BDNF-AS-DT* provide an additional layer of regulation over BDNF signaling during development and in response to neuronal activity, which are relevant in the etiopathogenesis of neuropsychiatric disorders.

## Results

### Characterization of novel divergent antisense transcription in the human BDNF locus

Novel transcription and splicing events can be pinpointed in RNAseq data by identifying reads that do not align to known transcript sequences. We re-analyzed a large RNAseq dataset from postmortem dorsolateral prefrontal cortex (DLPFC) tissue from neurotypical control donors and individuals diagnosed with schizophrenia (*29*). RNAseq libraries in this dataset were generated using Poly-A^+^ selection, so stable transcripts should be more readily detected compared with unstable transcripts (e.g. short bidirectional transcripts, upstream antisense RNAs or promoter upstream transcripts) (*30*). To comprehensively characterize the complex gene regulation of *BDNF*, we used the Integrative Genomics Viewer software (IGV) (*31, 32*) to identify novel transcription and splicing events. Analyzing this dataset (*29*) (see **Table S1** for sample characteristics), we noted reads in the 5’ upstream region of *BDNF* exon *I* (**Fig.1**) that are not annotated in the current reference transcriptome (Ensembl build GRCh38/hg38). These reads aligned to the positive DNA strand, antisense to *BDNF*, which is transcribed from the negative strand (**Fig.1**). Some reads partially overlapped *BDNF* exons *I* and *II*; however, most transcripts were detected in the region 5’ to the *BDNF* exon *I* transcription start site (TSS). We validated existence of these reads with PCR on cDNA from independent DLPFC postmortem human brain samples, followed by cloning and sequencing (**Fig.S1**). We named this new gene *BDNF divergent transcript* (symbol: *BDNF-DT*), consistent with the recommendations of the HUGO human gene nomenclature committee (*33*). We used 5’ and 3’ RACE-PCR with gene-specific primers on the novel exons to detect the 5’ start and the 3’ end of *BDNF-DT* (**Fig.S1**). We found two alternative 3’ ends, located respectively 3,720 and 52,445 kb upstream of *BDNF* exon *I* and three alternative 5’ starts, which overlap, respectively, *BDNF* exon *I*, *BDNF* exons *II* and the exon *I* of *BDNF-AS*, the previously annotated antisense in the *BDNF* locus. After defining the 5’ starts and the 3’ ends, we performed end-to-end PCR to characterize the full-length structure of the divRNAs: we detected three main transcripts of *BDNF-DT* (here referred to as *BDNF-DT1*, *2*, and *3*) and one *BDNF-AS-DT* readthrough transcript (**Fig.1** and **Fig.S2**; see **Table S2** for transcript sequences), that is, a RNA molecule formed via the splicing of exons from *BDNF-AS* and *BDNF-DT*. Of note, *BDNF-AS-DT* does not overlap the protein-coding exon of *BDNF*, which differs from *BDNF-AS*. We also re-analyzed RNAseq data from an independent postmortem DLPFC dataset (*34*), as well as datasets from postmortem human hippocampus (*34, 35*) and anterior cingulate cortex (*36*) (**Fig.S3,** see **Table S1** for sample characteristics). Because RNAseq libraries in these datasets were generated using ribosomal RNA depletion rather than Poly-A+ selection, unstable transcripts may have been detected, likely increasing transcriptional noise (*30, 37, 38*). However, coverage of RNAseq reads in these data is consistent with the existence of *BDNF* divergent and readthrough transcription not only in DLPFC (**Fig.S3a,b**), but also in hippocampus (**Fig.S3c,d**), including within the dentate gyrus (**Fig.S3e,f**), as well as within the subgenual and dorsal subdivisions of the anterior cingulate cortex (**Fig.S3g,h**). Further, by exploring a comprehensive resource of RNAseq data from 54 non-diseases tissues (*39*), we detected RNAseq reads supporting divergent transcription in the *BDNF* locus in many brain areas, including amygdala, anterior cingulate cortex, cerebellum, frontal cortex, hippocampus, and hypothalamus. Among the non-brain tissues, divergent transcription was blunted in most tissues, but particularly high in the pituitary gland (**Fig.S3i**).

### Relationship between BDNF-DT, BDNF-AS-DT and BDNF transcripts in human cortex across development

To investigate how *BDNF-DT* expression changes in DLPFC across development we further examined the postmortem human Poly-A^+^ RNAseq dataset described above (*29*) (**Table S1**). Analyzing data from just neurotypical brain donors, *BDNF-DT* expression is associated with age (N=296, F=11.03, p=2.47e-06); it increases during prenatal and neonatal life, until approximately 1 year of age, slightly decreases during childhood, until 12 years old, rises during adolescence and young adulthood, until 25 years old, and then drops during adulthood and senescence (**Fig.2a**). These changes mirror the developmental trajectory of *BDNF* in early life and adulthood, where expression increases until early postnatal life, followed by a decrease at the childhood-to-adolescence transition, and a further decrease during adulthood and senescence (**Fig.2b**). The *BDNF-AS-DT* transcript is also developmentally regulated (N=296, F=7.582, p=8.08e-06) and its association with age is similar to that of *BDNF-DT* and *BDNF* (**Fig.2a,b**). Specifically, it increases in prenatal and neonatal life, decreases in the childhood-adolescence transition, and decreases in adulthood (**Fig.2c**). However, the developmental trajectory of *BDNF-DT*, *BDNF-AS-DT* and *BDNF* are different compared with the previously characterized *BDNF-AS* (**Fig.2d**); *BDNF-AS* increases in adulthood and senescence, consistent with established patterns of other antisense RNAs (*40, 41*).

**Fig. 2.**
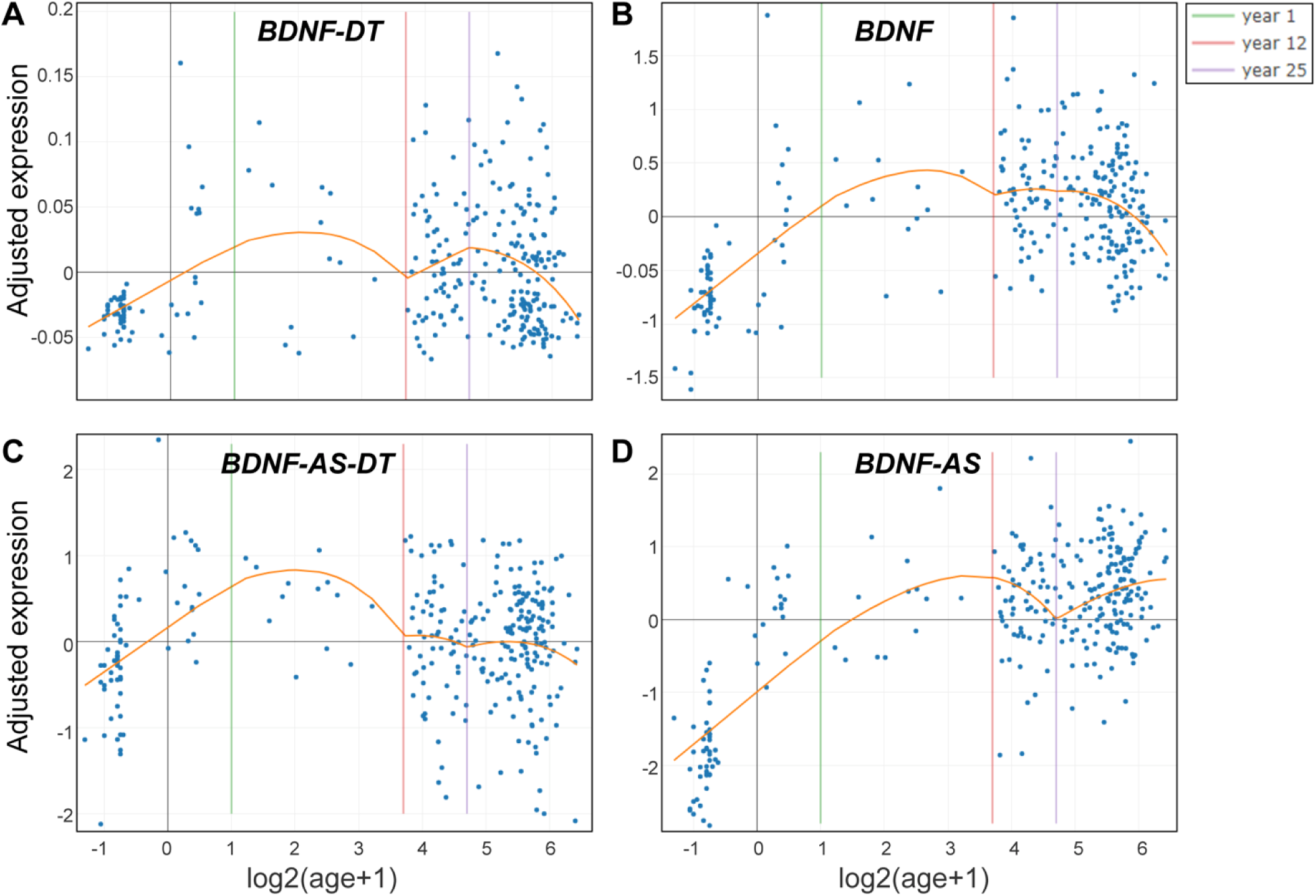
Developmental regulation of *BDNF-DT* and the known sense and antisense transcripts of the human *BDNF* locus. Adjusted gene expression (y-axis) of *BDNF-DT* (**A**), *BDNF* (**B**), *BDNF-AS-DT* (**C**), and *BDNF-AS* (**D**) is shown in relationship with age (orange line, x-axis, logarithmic scale). Age has a strong non-linear association with expression of *BDNF-DT* (N=296, F=11.03, p=2.47e-06), *BDNF* (N=296, F=41.91, p<2.2e-16), *BDNF-AS-DT* (N=296, F=7.58, p=8.08e-06), and *BDNF-AS* (N=296, F=132.92, p<2.2e-16).

We analyzed the relationship between *BDNF-DT*, *BDNF* and the other antisense genes in the subset of adult donors from the above RNAseq dataset (*29*) (age>13 years; **Table S1**). The expression of *BDNF-DT* is positively correlated with *BDNF* (N=219, r=0.168, t=2.768, p=0.006) although *BDNF* is highly expressed in some individuals with low *BDNF-DT* expression (**Fig.S4a**). *BDNF-DT* is not correlated with *BDNF-AS* (N=219, r=0.05, t=1.119, p=0.265; **Fig.S4b**), while it is weakly correlated with the readthrough transcript *BDNF-AS-DT* (N=219, r=0.10, t=2.038, p=0.0429; **Fig.S4c**). Of note, *BDNF-AS-DT* has a negative association with *BDNF* (N=219, r= −0.17, t= −2.109, p=0.036; **Fig.S4d**), and *BDNF-AS* (N=219, r= −0.178, t=-2.655, p=0.008; **Fig.S4e**). Finally, *BDNF* and *BDNF-AS* are not correlated (N=219, r= −0.017, t= - 0.056, p=0.956; **Fig.S4f**).

These associations suggest that *BDNF-DT*, *BDNF-AS*, and *BDNF-AS-DT*, are independently regulated lncRNAs with differing relationships to *BDNF*. While we did not detect an association between *BDNF-AS* and *BDNF*, we detected a positive correlation of *BDNF-DT* with *BDNF*, and a negative correlation between the *BDNF-AS-DT* readthrough transcript and *BDNF*. The similarity of the developmental trajectory of *BDNF-DT*, *BDNF-AS-DT* and *BDNF* supports the possibility that *BDNF-DT* and *BDNF-AS-DT* at least in part regulate BDNF, independently of *BDNF-AS*.

### Conservation of novel divergent antisense transcription in the mouse Bdnf locus

The *BDNF-DT* promoter shows high homology with the corresponding genomic sequence in the mouse genome. We extracted RNA from mouse frontal cortex and performed RT-PCR, to verify the existence of divergent transcription in the mouse *Bdnf* locus. We confirmed existence of an antisense amplicon connecting an exon partially overlapping exon *I* of *Bdnf* with an exon located in the region upstream the TSS of *Bdnf* exon *I*. We performed 5’- and 3’-RACE PCRs, and end-to-end PCR, confirming existence of the mouse *Bdnf-DT* gene. We detected four different transcripts, with high homology with the human *BDNF-DT1*, *2*, *3* (**Fig. S5, Table S3**). We did not find evidence for expression of a transcript homologous to the human *BDNF-AS* or the readthrough *BDNF-AS-DT* in the mouse genome. However, a mouse *Bdnf-AS* was recently described (*42*), which partially overlaps the *Bdnf-DT* gene. Based on our RACE results, we propose that the transcripts of the previously detected mouse *Bdnf-AS* are part of the *Bdnf-DT* gene, which has a further upstream 5’ exon, and a more complex structure (**Fig. S5**). Consistent with the recommendations of the HUGO human gene nomenclature committee (*33*), we propose that the mouse *Bdnf-AS* should be renamed *Bdnf-DT* for the following reasons: i) it is not antisense to the genomic span of the protein-coding portion of *Bdnf* (differently from the human *BDNF-AS*), ii) it does not show any homology with the human *BDNF-AS*, and iii) it is divergent to *Bdnf*, with which it shares at least one bidirectional promoter.

### Temporal dynamics of BDNF-DT and BDNF-AS-DT in response to neuronal depolarization

The positive correlation between *BDNF-DT* and *BDNF* expression across the lifespan (**Figs.2**, **S4**) supports the possibility of a regulatory role played by *BDNF-DT* with respect to *BDNF*. Because *BDNF* is strongly regulated by neuronal activity (*43–45*), we investigated whether *BDNF-DT* was also induced by neuronal depolarization. We first probed the relationship between KCl-induced depolarization and *Bdnf-DT* expression in mouse primary cortical cultures **(Fig.3)**. We observed a robust upregulation in *Bdnf I-IX* and *Bdnf II-IX* expression at 3 and 6 hours after stimulation with 100 mM KCl (**Fig.3a,b**). The highest level of expression, however, was observed after 6 hours of KCl stimulation followed by the wash out (WO) of KCl-containing media. *Bdnf I-IX* and *Bdnf II-IX* expression peaked ∼300 and ∼50 fold, respectively, at 3 hours post WO and gradually decreased to baseline over the next 24 hours (**Fig.3a,b**). *Bdnf IV-IX* displayed a different time-course of upregulation with a ∼ 30-fold induction at 3 hours, and a peak expression of nearly 60-fold at 6 hours after KCl stimulation with a gradual return to baseline after WO over the next 24 hours (**Fig.3c**). Interestingly, *Bdnf-DT* expression showed a temporal delay with respect to BDNF, in that it remained at baseline during the 6 hours of KCl stimulation, peaked 20-fold at 6 hours following WO, and returned to baseline over the next 24 hours (**Fig.3d**).

**Fig. 3.**
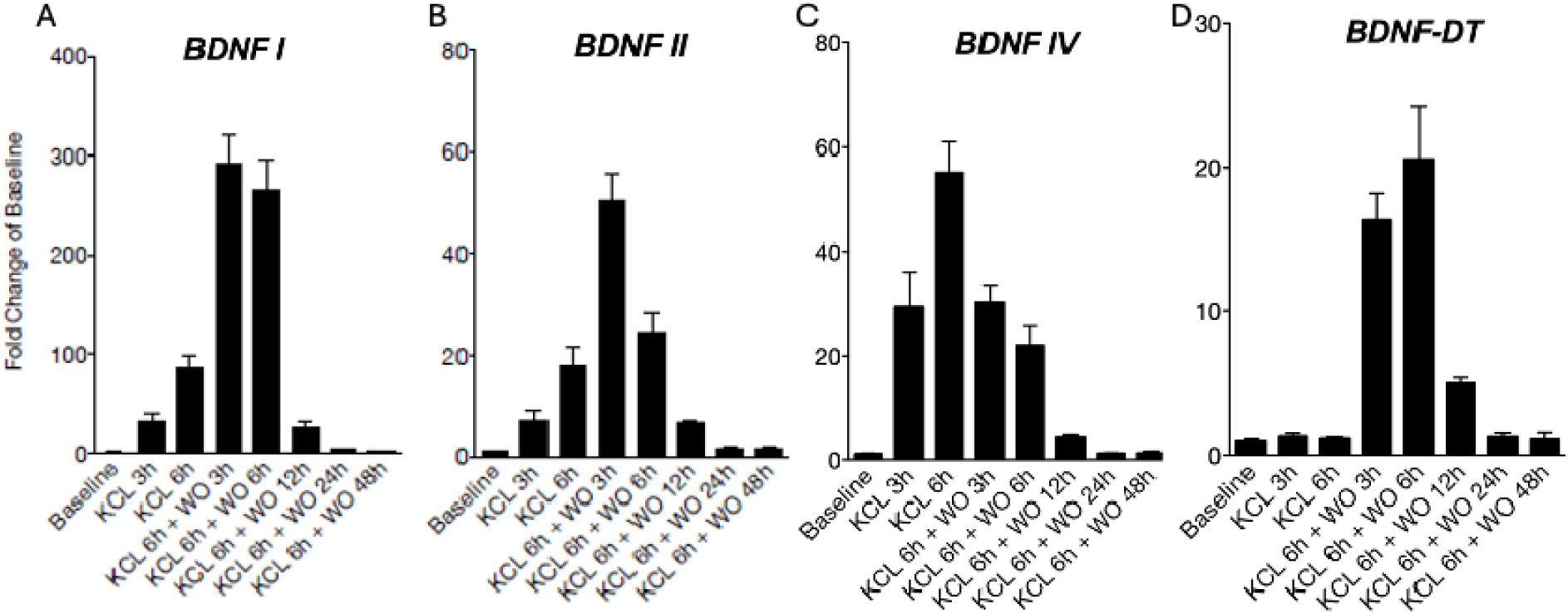
Temporal dynamics of *Bdnf* and *BDNF-DT* expression after depolarization in primary mouse cortical neurons. Expression of *Bdnf I-IX* transcript (**A**, *BDNF-I*, RefSeq: NM_007540.4), *Bdnf II-IX* transcript (**B**, *BDNF-II*, RefSeq: NM_001048139.1), *Bdnf IV-IX* transcript (**C**, *BDNF-IV*, RefSeq: NM_001048141.1), and *Bdnf-DT* (**D**) transcripts is shown at different time points after KCl-induced depolarization and wash-out (WO), in mouse cortical neurons. RNA expression, measured with qPCR, is represented as fold-change versus vehicle (baseline). KCl-induced neuronal depolarization causes an increase of the expression of all the *Bdnf* transcripts, and of *Bdnf-DT*, but with different temporal dynamics. RNA expression, measured with qPCR, is represented as fold change vs. Baseline. All data are expressed as mean□±□standard error of the mean (SEM).

We then examined *BDNF*, *BDNF-DT* and *BDNF-AS-DT* expression in response to induced depolarization in an immortalized human neural progenitor cell (NPC) line, which is derived from the ventral mesencephalon and can be differentiated into neurons or glia, ReNcell VM (*46*). Here, we quantified expression of *BDNF IX* (pan *BDNF* encompassing all coding transcript variants), *BDNF IV-IX*, *BDNF-DT*, and *BDNF-AS-DT* at different time points: 6 hours after depolarization alone or 6 hours after depolarization followed by 3, 6, 12, and 24 hours after WO (**Fig.4**). *BDNF*-*IX* and *BDNF IV-IX* peaked after 6 hours of stimulation, followed by 3 hours after WO, 3-fold and 8-fold, respectively, and returned to baseline over the next 24 hours (**Fig.4a,b**). We observed a strong effect of KCl-induced depolarization on *BDNF-DT*, an 11-fold increase after 6 hours of stimulation followed by 3 hours after WO, with a return to baseline over the next 24 hours (**Fig.4c**). The *BDNF-AS-DT* readthrough transcript was also elevated after depolarization, although less strongly compared with *BDNF-DT* (**Fig.4d**). These data support the hypothesis that *BDNF-DT* and *BDNF-AS-DT* are co-regulated or play a regulatory role with respect to *BDNF*, as their expression peaks simultaneously or follows *BDNF*’s upregulation both in mouse and human KCl-depolarized cell cultures.

**Fig. 4.**
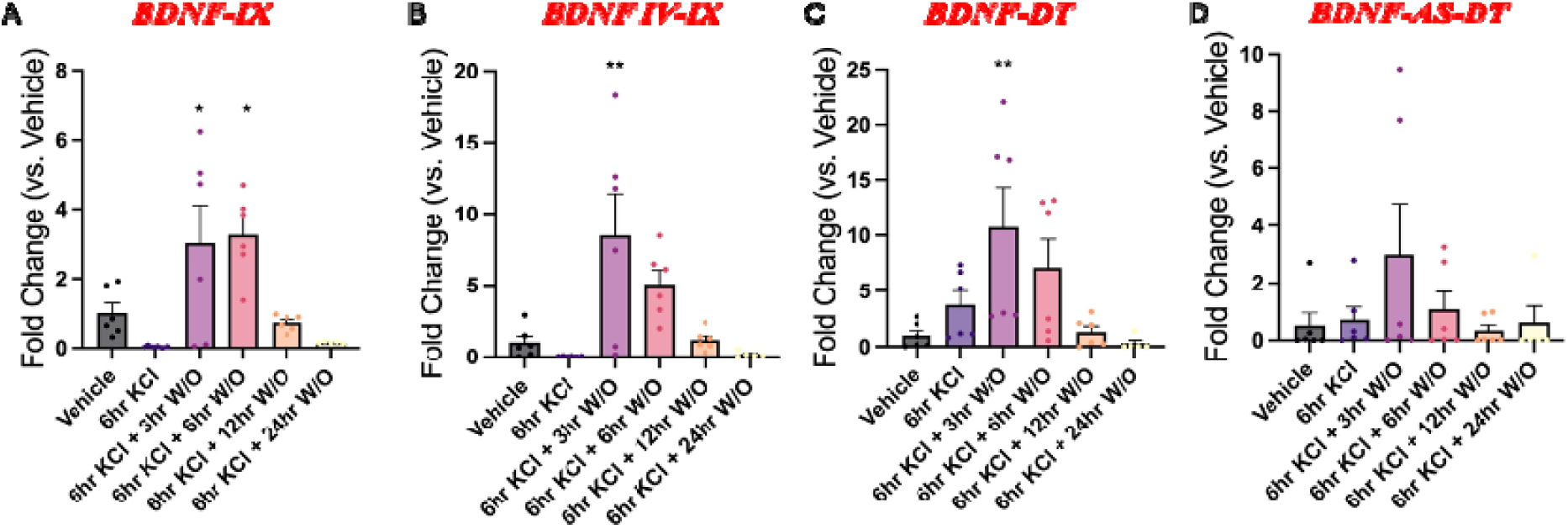
Temporal dynamics of *BDNF*, *BDNF-DT*, *BDNF-AS-DT* transcript expression after depolarization in the human ReNcell VM neural progenitor cells. Expression of total *BDNF* mRNA (**A**, *BDNF-IX*), the *BDNF* exon IV containing transcript (**B**, BDNF IV-IX, RefSeq: NM_001048141.1), *BDNF-DT* (**C**), and *BDNF-AS-DT* readthrough transcript (**D**) is shown at different time-points after KCl-induced depolarization, in human ReNcell VM neural progenitor cells. RNA expression, measured with qPCR, is represented as fold-change vs. vehicle. KCl-induced neuronal depolarization causes an increase of the expression of *BDNF*, *BDNF-DT*, and *BDNF-AS-DT*. RNA expression, measured with qPCR, is represented as fold-change vs. Vehicle. All data are expressed as mean□±□standard error of the mean (SEM). Individual comparisons; *p□<□0.05, **p□<□0.01.

### CRISPR-mediated regulation of BDNF induces BDNF-DT in human neural progenitor cells

The ReNcell VM line has previously been used for CRISPR gene editing and *BDNF* gene expression studies (*47*). To functionally test whether targeted upregulation of *BDNF* leads to an increase in *BDNF-DT* expression, we engineered a novel ReNcell VM NPC line to constitutively express a CRISPR activation (CRISPRa) system for gene upregulation (**Fig.S6a**). ReNcell VM cultures were transduced with high titer lentivirus carrying deactivated Cas9 (dCas9) endonuclease fused to a strong transcriptional activator, VP64-p65-Rta, (VPR) (*45*). Cells expressing VPR were propagated to create a stably transduced VPR-ReNcell VM line, which, like the traditional ReNcell VM line can be differentiated into neurons (**Fig.S6b**). Stable transduction was confirmed with single molecule *in situ* hybridization (smFISH) for Cas9 (**Fig.S6c**) and neural activity was verified with electrophysiological recordings (**Fig.S6d-e**).

To manipulate the expression of individual *BDNF* transcript variants, VPR-ReNcell VM cultures were transduced with lentiviruses carrying single guide RNAs (sgRNAs) targeting individual *BDNF* promoters at coding exons *I* or *IV* (**Table S4**). Previous work in cultured rat neurons and rat hippocampus validated the efficiency and selectivity of this CRISPRa system (*45*). Transduction of VPR-ReNcell VM line with *sgBDNF I* resulted in a 30-fold increase of *BDNF I-IX* (**Fig.5a**), but not *BDNF IV-IX* expression (**Fig.5b**). Transduction with *sgBDNF IV* resulted in an almost 800-fold increase of *BDNF IV-IX* (**Fig.5b**), but not *BDNF I-IX* expression (**Fig.5c**). CRISPRa-mediated upregulation of *BDNF I-IX* and *BDNF IV-IX* corresponded to >2-fold and to >3-fold significant increase of *BDNF-DT* (**Fig.5c**). These results indicate that the induction of the expression of BDNF-protein coding transcripts also causes an increase of *BDNF-DT*. *BDNF-AS* and *BDNF-AS-DT* were not detectable in this in vitro system.

**Fig. 5:**
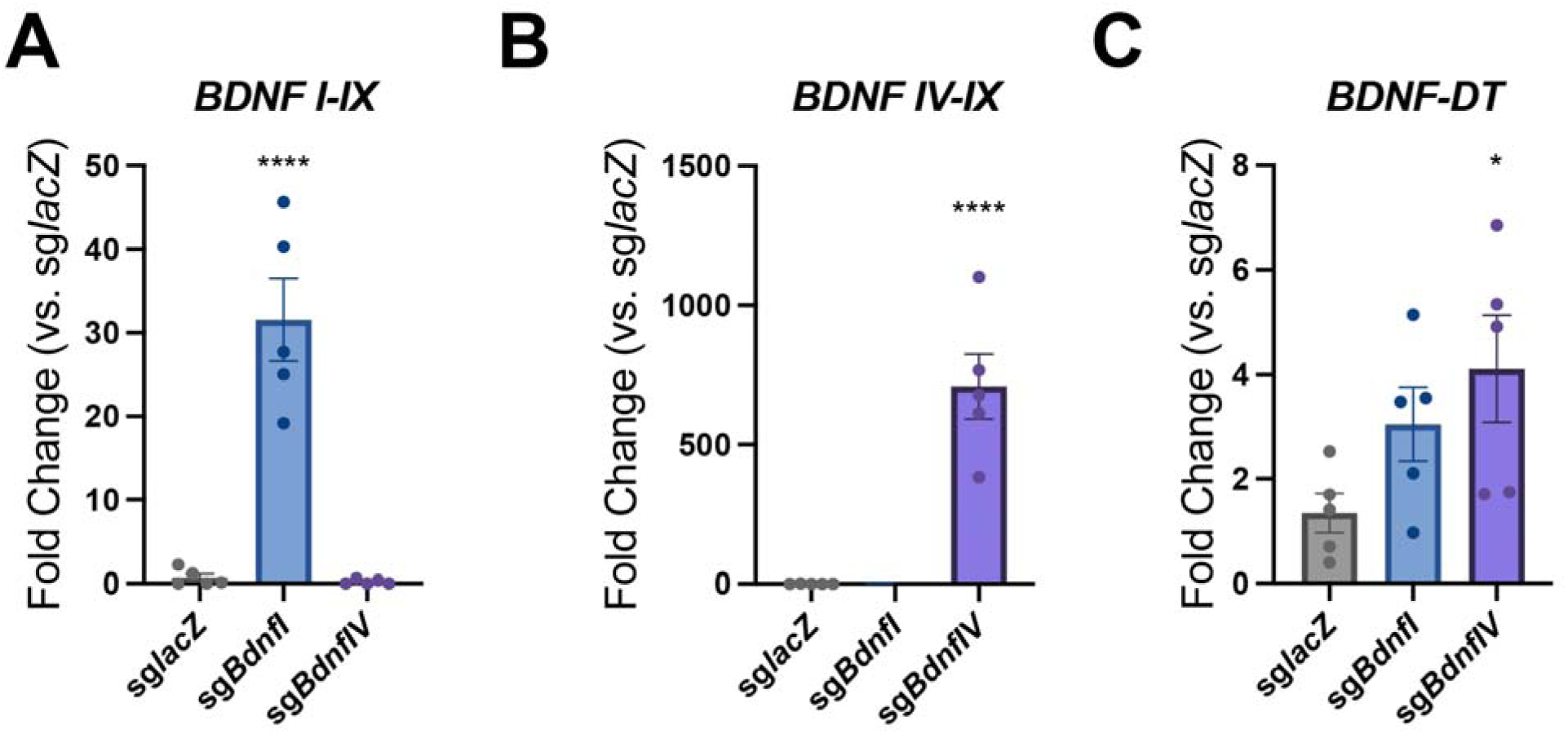
Upregulation of *BDNF-DT* using CRISPRa in VPR-ReNcell VM culture. Expression of *BDNF I-IX* (**A**), *BDNF IV-IX* (**B**) and *BDNF-DT* (**C**) transcripts is shown after CRISPRa of *BDNF I* promoter (sg*Bdnf I*) and of *BDNF IV* promoter (sg*Bdnf IV*). Activation of *BDNF I* promoter induces a 30-fold increase of *BDNF* I-IX transcript (**A**), a 2-fold increase of *BDNF-DT* (**C**), and has no off-target effect on *BDNF* IV-IX transcript (**B**). Activation of *BDNF IV* promoter induces a 800-fold increase of the *BDNF* IV-IX transcript (**B**), a 3-fold increase of *BDNF-DT* (**C**), and has no off-target effect on the *BDNF* I-IX transcript (**A**). RNA expression, measured with qPCR, is represented as fold-change vs. non-targeting control sgRNA, *lacZ*. All data are expressed as mean□±□standard error of the mean (SEM). Individual comparisons; *p□<□0.05, ****p□<□0.0001.

### Link between BDNF-DT, BDNF-AS-DT and schizophrenia

Our data demonstrate that *BDNF-DT* and BDNF-AS-DT are developmentally regulated, sensitive to neuronal activity, and can potentially regulate *BDNF*. We next analyzed a relationship between these novel genes and schizophrenia, a neurodevelopmental disorder linked to alterations in neuronal activation (*48*) and with a GWAS significant association in the *BDNF* locus (*49*). Leveraging the postmortem human RNAseq dataset described above (*29*), we investigated expression of *BDNF-DT* and the *BDNF-AS-DT* readthrough transcript in DLPFC tissue from control brain donors versus schizophrenia. While expression of *BDNF-DT* is significantly higher in donors with schizophrenia than in neurotypical controls (N=378, t=3.805, p= 0.000167; **Fig.6a**), *BDNF-AS-DT* (N=378, t=-0.875, p= 0.3824), *BDNF* (N=378, t=0.572, p= 0.5675), and *BDNF-AS* (N=378, t=0.10, p= 0.920720) remain unchanged.

**Fig. 6:**
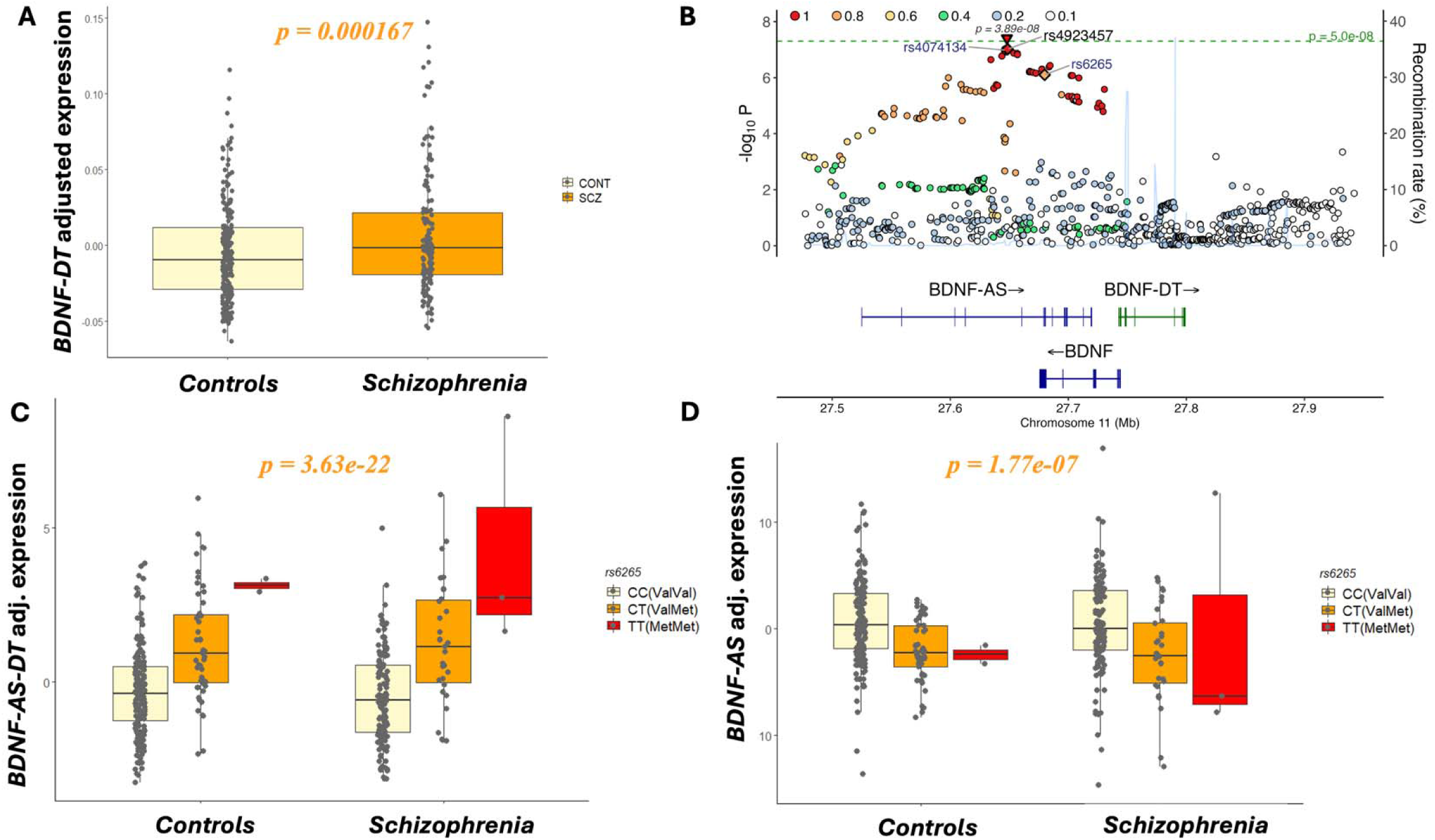
Divergent transcription, schizophrenia, and rs6265. **A)** Expression of *BDNF-DT* (adjusted values, y-axis) is higher in patients affected with schizophrenia (orange, right box on the x-axis) compared with in neurotypical controls (light yellow, right box on the x-axis); p-value is shown at the top. **B)** Schizophrenia GWAS plot visualization of the *BDNF* locus. The SNP rs49234957, indicated with two inverted triangles, is the variant with the most significant association with schizophrenia in the *BDNF* locus, and it reaches GWAS significance in the combined replication and discovery sample (upper triangle, p=3.89e-08). Rs49234957 is in high linkage disequilibrium with rs6265. **C)** Expression of the *BDNF-AS-DT* readthrough transcript (y-axis) is associated with the rs6265 genotype so that it increases with the number of T/Met alleles: specifically, it is higher in the TT/MetMet genotype (red box) and in the CT/ValMet (orange box) compared with the CC/ValVal (light yellow box), in both neurotypical controls (left, x-axis) and patients affected with schizophrenia (right, x-axis); p-value is shown at the top. **D)** Expression of *BDNF-AS* (y-axis) is associated with the rs6265 genotype so that it decreases with the number of T/Met alleles: specifically, it is lower in the TT/MetMet genotype (red box) and in the CT/ValMet (orange box) compared with the CC/ValVal (light yellow box), in both neurotypical controls (left, x-axis) and patients affected with schizophrenia (right, x-axis); p-value is shown at the top.

Because previous work suggests that differentially expressed genes in postmortem human brain tissue studies of schizophrenia are more likely reflecting consequences of illness rather than underlying genetic causes(*34, 50*), we leveraged Summary-based Mendelian Randomization (SMR) (*51*) to investigate whether genetic variation in the *BDNF* locus might be mediating schizophrenia risk through regulating expression of these transcripts. Of note, the most recent GWAS(*49*) identified the rs4923457 A/T single nucleotide polymorphism (SNP) as the genetic variant with the most significant association with schizophrenia in the *BDNF* locus. This SNP reaches GWAS-significance in the combined discovery and replication samples (p*_discovery-sample_*=9.52e-08, p*_combined-sample_*=3.89e-08, OR for A allele=1.05; **Fig.6b**) but, because its alleles are complementary base pairs, it is not suitable as an instrumental variable for the SMR analysis, given the risk of potential inconsistent strand alignment across different DNA genotyping batches. Of note, rs4923457 is in high LD (D’=0.955, R^2^= 0.798) with rs6265 (also known as BDNF Val66Met), a highly studied functional variant in psychiatric genetics(*52*), which lies in the BDNF pro-region domain and produces a non-synonymous substitution at codon 66: C → T/Valine → Methionine (Val66Met). This SNP is directly linked to subcellular trafficking of BDNF, and is associated with behavior in both human and mice: in particular, the Met allele has been shown to disrupt the activity-dependent secretion of BDNF by inhibiting sorting of BDNF into dense-core vesicles (*52*). However, GWAS data indicate that the T allele (Met-BDNF), correlated with the rs4923457 T allele, is associated with decreased risk for schizophrenia [odds ratio (OR) = 0.96 and 0.95, respectively for rs6265 T allele and rs4923457 T allele]. We therefore performed SMR with rs6265 as the instrumental variable, and we found that the readthrough transcript *BDNF-AS-DT* has the most significant potential causal association with schizophrenia, so that lower *BDNF-AS-DT* genetically predicted expression is associated with the disease (p=9.89e-06 in the whole sample and p=7.64e-05 in the European ancestry sample; **Table S5**). SMR also detected an association with opposite sign between *BDNF-AS* and schizophrenia, albeit less significantly compared with *BDNF-AS-DT* (**Table S5**). Results were consistent when running SMR without pre-selecting any candidate SNP (**Table S6**). Indeed, the eQTL analysis shows that rs6265 is significantly associated with the expression of *BDNF-AS-DT* (N=378, t=10.365, p=3.63e-22; **Fig.6c).** The relationship of rs6265 with *BDNF-AS-DT* is stronger compared with rs4923457 (N=378, t=6.829, p=3.72e-11), the top schizophrenia GWAS-index SNP in this locus. This association of rs6265 with *BDNF-AS-DT* is present both in neurotypical controls (N=219, t=7.267, p=8.38e-12) and in patients with schizophrenia (N=159, t=7.073, p=7.11e-11; **Fig.6c**).

We also analyzed the relationship of rs6265 with expression of the other genes in the same locus. While we did not detect any significant association of rs6265 with *BDNF-DT* (N=378, t=0.608, p=0.54) and *BDNF* (N=378, t=0.802, p=0.42), we found a significant association between rs6265 and *BDNF-AS*, with opposite directionality compared with the *BDNF-AS-DT* (N=378, t= −5.328, p=1.77e-07; **Fig.6d**). In other words, the T allele (Met-BDNF) is associated with higher expression of *BDNF-AS-DT* and lower expression of *BDNF-AS*. Because the *BDNF-AS-DT* transcripts differs from *BDNF-AS* not only for the link with *BDNF-DT,* but also for the skipping of an exon common to all the *BDNF-AS* transcripts (hg38: chr11:27,658,240-27,658,462), and containing rs6265 (hg38: chr11: 27,658,369), these findings suggest that rs6265 genotype may also be associated with an effect on transcriptional splicing. In particular, the rs6265 T allele appears to be associated with the skipping of this exon common to all the *BDNF-AS* transcripts, leading to opposite effect on expression of *BDNF-AS-DT* (increase) and *BDNF-AS* (decrease).

Intragenic DNA methylation has been associated with exon inclusion during transcription (*53, 54*), thus we explored the possible role of DNA methylation in the relationship between rs6265 and *BDNF-AS-DT* expression. Because the rs6265 C>T substitution abolishes a CpG site in the position chr11:27658369, we analyzed the relationship between methylation of this CpG site and *BDNF-AS-DT* expression, leveraging DNA methylation bisulfite sequencing DLPFC data available in a sample subset (N=141, **Table S1**). We detected a negative relationship between DNA methylation at this site and *BDNF-AS-DT* expression (N= 141, Z= −3.86, p= 0.0001). By extending the analyses to other CpGs in the same locus, we note that this CpG in position chr11:27658369 is the top methylation site associated with expression of *BDNF-AS-DT* (**Table S7**). As predicted, the rs6265 C(G)/T(G) SNP is associated with methylation of the CpG at position chr11:27658369 (N= 141, t= −19.31, p=4.84e-39; **Fig.S7**), consistent with previous work (*55*).

### Co-expression gene networks associated with BDNF, BDNF-DT and BDNF-AS-DT in the human cortex

To explore convergence and divergence in expression patterns linked with *BDNF-DT*, *BDNF-AS-DT*, *BDNF*, and *BDNF-AS*, we performed brain coexpression network analyses using each of the above as seed genes, in the subset of neurotypical controls (N=318) from the above referenced DLPFC RNAseq dataset (*29*) (**Fig. 7a-d**, **Fig.S8**, and **Tables S8-11**). Genes significantly co-expressed with *BDNF-DT* were associated with neurogenesis and synaptic function, and with serotonin, dopamine, and opioid signaling (**Fig. 7d**, **Fig.S8a**, and **Table S8a**); indeed, these genes were associated with activation of BDNF and inhibition of REST, the neuron-restrictive silencing factor (also known as NRSF), a fundamental transcription repressor element involved in suppressing expression of neural genes (*56–58*) (**Table S8b**). Genes significantly co-expressed with *BDNF* were also associated with neurogenesis and synaptic function, and inhibition of REST (**Fig.S8b**, and **Table S9a,b**). On the other hand, genes significantly co-expressed with *BDNF-AS* were associated with inhibition of synaptogenesis and dopamine pathways, and REST activation (**Fig.S8c**, and **Table S10a,b**), while genes co-expressed with *BDNF-AS-DT* were linked with protein ubiquitination, and *SOX2A* activation (**Fig.S8d**, and **Table S11a,b**). Of note, the ubiquitin proteasome pathway is linked to *BDNF* expression (*45*), and can impact transcription termination by RNA polymerase II (*59*), which is directly related to readthrough transcription under stress conditions (*15–17*).

**Fig. 7:**
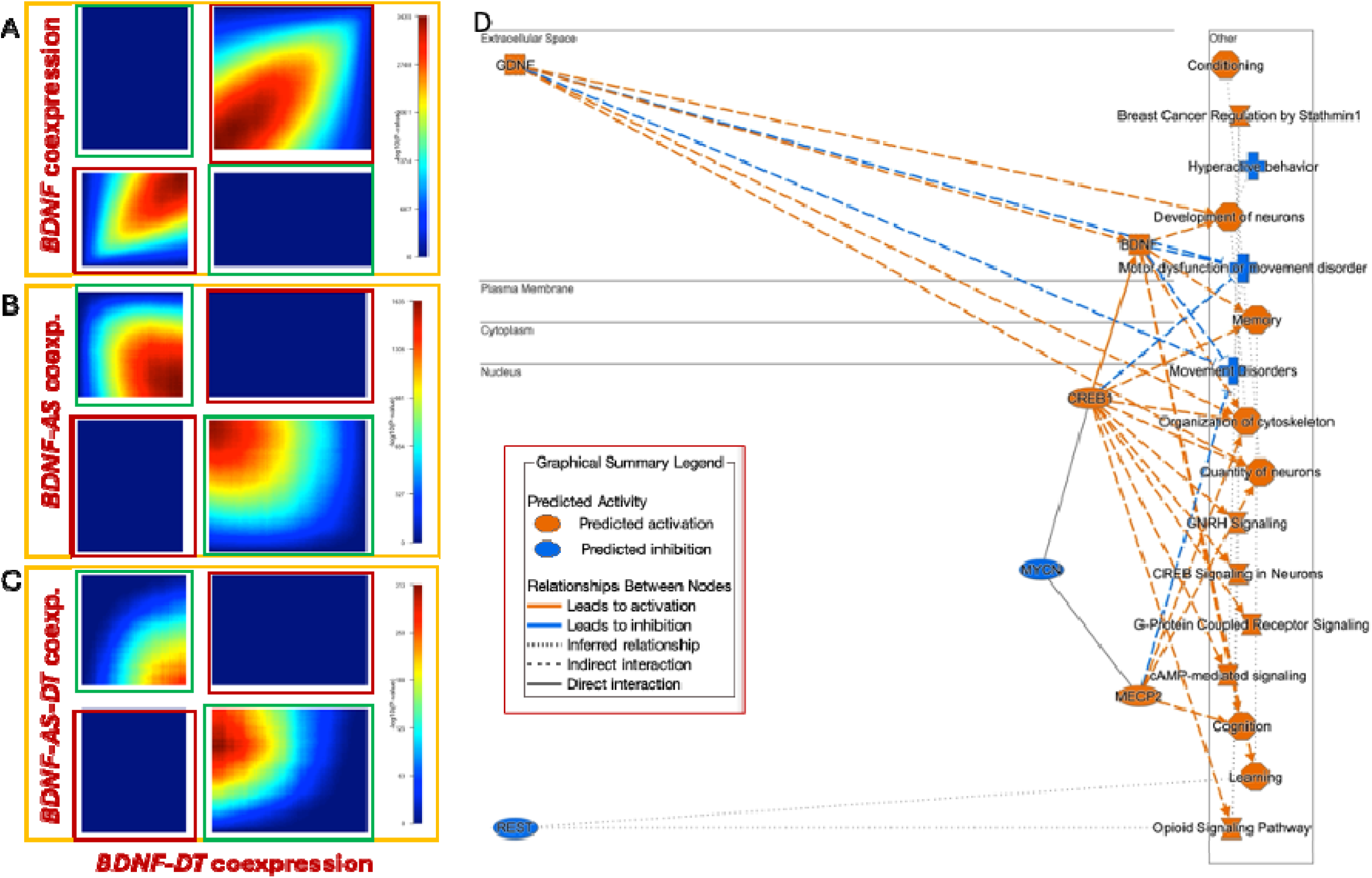
Coexpression gene networks associated with *BDNF-DT*, *BDNF-AS-DT*, *BDNF-AS*, and *BDNF*, in human DLPFC. **A-C)** RRHO2 heatmaps showing concordant (bottom-left and top-right quadrant in each panel, highlighted in a red frame) and discordant (top-left and bottom-right quadrant in each panel, highlighted in a green frame) association of *BDNF-DT* coexpression network (x-axis, **A-C**) with coexpression network of *BDNF* (y-axis, **A**), *BDNF-AS* (y-axis, **B**), *BDNF-AS-DT* (y-axis, **C**). Color bars represent negative logarithm of p value of the overlap from the hypergeometric test. *BDNF-DT* show a concordant pattern of association with gene expression compared with *BDNF* (**A**), and discordant pattern of association compared with *BDNF-AS* (**B**), and *BDNF-AS-DT* (**C**). **D**) Ingenuity Pathway Analysis summary graph with subcellular localization of the activators (orange) and inhibitors (blue) upstream regulators of the *BDNF-DT* gene coexpression network, and visualization of their effect on pathways predicted to be activated (orange) and inhibited (blue) in presence of the gene expression changes associated with the *BDNF-DT* gene coexpression network. Legend in the red box.

We also used a threshold-free algorithm (*60*) to detect concordant and discordant patterns of associations with potential biological relevance of *BDNF-DT* with *BDNF*, *BDNF-AS*, *BDNF-AS-DT*. Briefly, we analyzed whether the genes co-expressed with *BDNF-DT* were co-expressed with *BDNF*, *BDNF-AS*, *BDNF-AS-DT* with the same directionality (positive vs. positive, and negative vs. negative: concordant pattern) or with opposite directionality (positive vs. negative, or negative vs. positive: discordant pattern). We detected a highly concordant pattern of associations of gene expression between *BDNF-DT* and *BDNF* (**Fig.7a**), while we detected discordant pattern of association of gene co-expression of *BDNF-DT* with *BDNF-AS* (**Fig.7b**), and less strongly with *BDNF-AS-DT* (**Fig.7c**). These results provide additional evidence that *BDNF* and *BDNF-DT* share similar expression patterns, while their relationship with co-expressed genes contrasts with that of *BDNF-AS* and the *BDNF-AS-DT* readthrough transcript.

## Discussion

RNAseq in human brain has revealed many novel transcripts and isoforms of known genes. In addition, it is now understood that transcription occurs from non-coding regions of the genome. Divergent transcription from bidirectional promoters substantially contributes to overall levels of transcription, and it has been proposed that the majority of mammalian lncRNAs originate from divergent transcription of gene pairs (*19*). Next generation sequencing also indicates that readthrough transcription is a common phenomenon (*17*). However, it is still not known whether divergent and readthrough transcription are functionally relevant.

We performed a genomic characterization of a divRNA in the human and mouse *BDNF* locus, which we named *BDNF-DT*. Consistent with other divRNAs, our data suggest that *BDNF-DT* is evolutionary conserved (*61*). *BDNF-DT* transcription start sites overlap known *BDNF* exons, they are transcribed antisense to *BDNF*, and they have a 3’ end located 5’ upstream of *BDNF* exon *I* (**Fig.1**). CpG islands overlap the transcription start site of *BDNF-DT* (**Fig.S2**, **S5**), and the 5’ region of *BDNF-DT* overlaps areas of *BDNF* exon *II* that bind the RE1-Silencing Transcription factor (REST) (*56–58, 62*). The REST binding site (neuron-restrictive silencer element, NRSE) is located on the opposite strand of the *BDNF* gene, downstream of the 5’ region of *BDNF-DT.* This is consistent with the possibility that REST regulates directly *BDNF-DT* expression, which may subsequently regulate *BDNF*. Indeed, our co-expression network analyses indicate that genes co-expressed with *BDNF* and *BDNF-DT* are predicted to be regulated by REST (**Fig.7d**, **S8a,b**).

In addition, we detected *BDNF-DT* isoforms spliced with the known *BDNF-AS*, which constitute novel *BDNF-AS-DT* readthrough transcripts. *BDNF-DT* and its readthrough transcript are developmentally regulated - their expression increases starting from prenatal and neonatal life, decreases in the transition from childhood to adolescence, peaks again around 25 years of age and then decreases (**Fig.2**). These changes in expression across development mirror those of *BDNF*; while on the other hand, they differ from that of known *BDNF-AS*, whose expression increases during adulthood. While *BDNF-DT* and *BDNF* expression patterns are positively correlated, we detected a negative correlation between *BDNF-AS-DT* and *BDNF*, and no correlation between *BDNF-AS* and *BDNF* (**Fig.S4**). These findings suggest that *BDNF-DT* and *BDNF-AS-DT* are expressed in similar developmental contexts as *BDNF*, but they may regulate *BDNF* in contrast to *BDNF-AS*. Indeed, gene coexpression networks revealed highly concordant patterns of associations between *BDNF-DT* and *BDNF* (**Fig.7a**), and discordant patterns between *BDNF-DT* and *BDNF-AS* (**Fig.7b**) or *BDNF-DT* and *BDNF-AS-DT* (**Fig.7c**).

*BDNF*, *BDNF-DT* and *BDNF-AS-DT* expression increases in response to KCl-induced depolarization (**Fig.3**, **4**). In cultured mouse neurons we observed differing temporal dynamics in the expression patterns between activity-induced *Bdnf* and *Bdnf-DT*. Specifically, the increase of *Bdnf* transcripts precedes the increase of *Bdnf-DT*, which occurs simultaneously with a decrease of the *Bdnf* coding transcript. This evidence raises the possibility that *BDNF-DT* plays a role in regulating *BDNF* levels, via an autoregulatory negative feedback loop. Supporting this idea, CRISPR-mediated induction of *BDNF* exon *I* and *IV* transcription in human NPCs drives subsequent *BDNF-DT* expression (**Fig.5c**). These data raise the possibility that the divRNAs may regulate the optimal ‘balance’ of *BDNF*. While *BDNF* expression is essential for synaptic plasticity and trophic support, its levels must be tightly controlled to prevent hyper-excitability(*24–28*). We therefore propose that *BDNF-DT* and *BDNF-AS-DT* are part of a regulatory mechanism, where expression of *BDNF* induces expression of *BDNF-DT*, which is required for the splicing of *BDNF-AS* with *BDNF-DT*, thus originating *BDNF-AS-DT*. *BDNF-AS-DT* may therefore represent a key regulator of BDNF. Additional mechanistic experiments will help to definitively test this hypothesis, which is consistent with our data showing an effect of CRISPR mediated *BDNF* transcription on *BDNF-DT* expression (**Fig.5c**), and an effect of KCl-induced depolarization on the expression, first of *BDNF*, and then of *BDNF-DT* and *BDNF-AS-DT* (**Fig.4**). Further, this hypothesis is consistent with our observations of a positive correlation of *BDNF-DT* with *BDNF* and *BDNF-AS-DT*, which is negatively correlated with *BDNF* (**Fig.S4**).

Schizophrenia is a neurodevelopmental disorder that is linked with altered *BDNF* signaling (*20, 21, 63*) and neuronal activity (*48*). Hence, we reasoned that the developmental regulation of *BDNF-DT* and *BDNF-AS-DT*, and their potential ability to regulate *BDNF* in response to neuronal activity, may contribute to established links between schizophrenia and BDNF signaling. To probe this hypothesis, we investigated whether *BDNF-DT* and *BDNF-AS-DT* were differentially expressed in patients affected with schizophrenia compared with neurotypical controls, and whether their expression was associated with genetic risk for schizophrenia in the *BDNF* locus. We observed higher expression of *BDNF-DT* in the DLPFC of patients with schizophrenia, compared with controls, and no significant differences in *BDNF-AS-DT* expression (**Fig.6a**). These findings should be interpreted with caution since differences in gene expression between patients and controls tend to be confounded by medication, comorbid health issues and socioeconomic factors. Hence, even if observed differences are robust, they may reflect consequences of the disorder, rather than risk or cause. Genetic association, in contrast, is independent of illness state and confounders. In this context, SMR indicates a significant association of genetically predicted expression of *BDNF-AS-DT* with schizophrenia. *BDNF-AS-DT* is indeed associated with rs6265, a SNP in the BDNF locus that shows association with schizophrenia in the current GWAS. Rs6265 is in strong linkage disequilibrium with the SNP showing the most significant association with schizophrenia in the *BDNF* locus (*49*). The association is highly significant in the whole sample and, separately, both in patients with schizophrenia and controls. In particular, the C/Val-coding and risk-associated allele is associated with lower expression of *BDNF-AS-DT* (**Fig. 6c**) and higher expression of the canonical *BDNF-AS* gene (**Fig. 6d**).

Current data about rs6265 may appear potentially paradoxical, given previous allele-specific findings showing deleterious effects of the T/met-coding allele on diverse brain phenotypes, and the association of the C/Val-coding allele with higher risk for schizophrenia. Our results suggest that the effects of the rs6265 val66met variant on BDNF activity and on the expression of the readthrough transcript are two distinct mechanisms, that may impact BDNF biology in a context specific manner. The effects of the C/Val-coding allele on activity-dependent secretion of BDNF and on lower expression of *BDNF-AS-DT* may converge, in specific developmental stages, on increasing the activity and the expression of BDNF, creating an excess of BDNF which may disrupt brain development, increasing risk for schizophrenia. At the cellular level, the Met allele has been shown to disrupt activity-dependent secretion of BDNF by inhibiting sorting of BDNF into dense core vesicles (*52*). Experimental studies have shown that the Met-BDNF results in an approximate 18% decrease in activity-dependent secretion in transfected cells carrying one Met allele and a 29% decrease in those transfected with two Met alleles (*64*). However, what is often neglected is that rs6265 lies not only in the BDNF pro-region domain but also in an exon common to all the *BDNF-AS* isoforms (hg38: chr11:27,658,240-27,658,462) which our findings show is not part of the *BDNF-AS-DT* readthrough transcript. An effect of rs6265 on the splicing of such exon is plausible, consistent with the fact that splice regulatory elements can lie within alternative exons and can be disrupted by sequence variations (*65*); in this regard, rs6265 may affect splicing by influencing the binding of transcription factors on the positive strand of the DNA, from which *BDNF-AS* and *BDNF-AS-DT* are transcribed. DNA methylation may contribute to mediate these effects, consistently with the relationship between rs6265 and intragenic methylation (**Fig. S7**), and the relationship between intragenic DNA methylation and exon inclusion during transcription (*53, 54*). Indeed, we show that the C/Val-coding allele is associated with higher methylation, lower expression of *BDNF-AS-DT* and higher expression of the canonical *BDNF-AS* gene, which incorporates the exon missing in the readthrough transcript. The C/Val-coding allele may therefore be associated not only with higher activity-dependent secretion of the BDNF protein but also with blunted expression of *BDNF-AS-DT*, with a possible impairment in the regulation of *BDNF* and a susceptibility to a detrimental excess of BDNF which may lead to hyperexcitability during early development, and higher risk for schizophrenia. This aspect of rs6265 biology is likely overlooked by most mechanistical studies on the SNP, in cellular and animal models, which are often based on the ectopic expression of constructs expressing the Val- or the Met-BDNF in a genomic context that does not include the *BDNF-AS*, *BDNF-AS-DT*, and *BDNF-DT*. Even if we cannot exclude the possibility that the relationship between rs6265 and *BDNF-AS-DT* expression is driven by other genetic variants in high LD with rs6265, the presence of such an association is still relevant to understand the clinical correlates of rs6265.

In conclusion, our study sheds a new light and poses new questions on the complex biology of *BDNF*, revealing that, in addition to the canonical *BDNF-AS* isoforms, divergent RNAs and readthrough transcripts linking *BDNF-AS* and the divRNAs may contribute to *BDNF* regulation. Moreover, our study provides novel insight on the mechanisms mediating or moderating the effect of the functional rs6265 polymorphism, one of the most studied SNPs in psychiatric genetics, raising the possibility that readthrough transcription contributes, directly or indirectly, to the effect of this functional polymorphism. Our study does not clarify whether divergent transcription and readthrough transcription represent just noise or phenomena with biological relevance. However, the evolutionary conservation, the developmental trajectory and the temporal dynamic after neuronal depolarization of *BDNF-DT* and *BDNF-AS-DT*, support a regulatory role on *BDNF*, potentially relevant for brain development and activity, and risk for schizophrenia.

## Materials and Methods

### Postmortem brain samples

Postmortem human brain tissue was obtained by autopsy primarily from the Offices of the Chief Medical Examiner of the District of Columbia and of the Commonwealth of Virginia, Northern District, all with informed consent from the legal next of kin (protocol 90-M-0142 approved by the NIMH/NIH Institutional Review Board). The National Institute of Child Health and Human Development Brain and Tissue Bank for Developmental Disorders (http://www.BTBank.org) provided additional postmortem prenatal, infant, child and adolescent brain tissue samples, by under contracts NO1-HD-4-3368 and NO1-HD-4-3383. The Institutional Review Board of the University of Maryland at Baltimore and the State of Maryland approved the protocol, and the tissue was donated to the Lieber Institute for Brain Development under the terms of a Material Transfer Agreement. Tissue acquisition, handling, processing, dissection, clinical characterization, diagnoses, neuropathological examinations, and quality control measures were performed following established protocols at the LIBD as previously described (*29, 66*).

### RNA extraction and sequencing

Postmortem tissue homogenates of dorsolateral prefrontal cortex gray matter (DLPFC) approximating Brodmann area 46/9 in postnatal samples and the corresponding region of PFC in prenatal samples were obtained from all subjects. Total RNA was extracted from ∼100 mg of tissue using the RNeasy kit (Qiagen) according to the manufacturer’s protocol. The poly(A)- containing RNA molecules were purified from 1 μg DNase-treated total RNA, and sequencing libraries were constructed using the Illumina TruSeq RNA Sample Preparation v2 kit. These products were then purified and enriched with PCR to create the final cDNA library for high-throughput sequencing using an Illumina HiSeq 2000 with paired-end 2 × 100 bp reads, as previously described (*29*). Poly(A)+ libraries include divRNAs, since these transcripts do have a Poly(A) tail, together with other features of mRNAs, such as the 7-methylguanosine cap, and being often subject to splicing (*19*). RNAseq data processing has been extensively previously described (*29*). Rs62625 and other genotypes were extracted from genotyping data obtained with HumanHap650Y_V3, Human 1M-Duo_V3, and Omni5 BeadChips (Illumina, San Diego, CA), as described in previous reference (*29*). Sample characteristics and demographics are summarized in **Table S1**.

### Pipeline for detection and validation of divRNAs

A similar procedure was performed to detect and validate divRNAs in the human *BDNF* and mouse *Bdnf* locus:

#### Detection of divergent transcription

Most divRNAs are not annotated in the reference transcriptome (GRCh38/hg38). Moreover, they are easily degraded, and they can have very low expression, so that transcripts assembly procedures are not completely reliable in detecting their expression and their structure. In addition, they can overlap exons of protein-coding genes, so that precise information about the expression of divRNAs cannot be obtained with measures that do not consider the directionality of transcription, such as exons and ‘expressed genomic regions (*67*)’, without defining their exact structure. For these reasons, we investigated the occurrence of divergent transcription at the promoter of protein-coding genes by visualizing RNAseq data with Integrative Genomics Viewer (IGV), a high-performance software for visualization and exploration of genomic data (*31*). After detecting sequencing reads in the promoter region of protein-coding genes, we confirmed their antisense direction by analyzing the consistence with the intron rules (*68*) of the genomic sequences overlapping the reads covering exon-exon splice junctions of the potential divRNAs.

#### PCR validation

We validated the novel sequences potentially belonging to the divRNAs with PCR. RNAs isolated from postmortem DLPFC were reverse transcribed using SuperScript First-Strand Synthesis System for RT-PCR (Invitrogen) to synthesize cDNA. Based on the RNAseq results, we designed primer pairs to amplify portions of the divRNAs, thus validating the novel exons detected with the RNAseq analysis (primer sequences available in **Table S4**).

#### RACE

5′ RACE and 3′ RACE were performed using SMARTer® RACE 5′/3′ Kit (Clontech) to determine the 5’ start and the 3’ end of divRNAs. RNAs isolated from postmortem human DLPFC were reverse transcribed to full-length cDNA using the reverse transcriptase and the oligo(dT)-anchor primer available in the kit, according to the manufacturer’s protocol (Clontech). The obtained cDNA was further amplified by a second PCR, using divRNA-specific primer, the PCR-anchor primer, and the Advantage Polymerase Mix, according to the manufacturer’s instruction (Clontech). Nested PCR was then performed on the RACE-amplicons, to confirm the presence of the divRNAs.

#### PacBio sequencing

PacBio sequencing was performed on the RACE amplicons where the nested PCR confirmed the presence of divRNAs. Three micrograms of purified PCR products were sequenced on an RSII instrument, according to the manufacturer’s protocols (Pacific Biosciences), by the Johns Hopkins Transcriptomics and Deep Sequencing Core Facility. Briefly, PCR products were purified twice by AMPure magnetic beads (Beckman-Coulter) to yield SMRTbell libraries. We size-selected PCR products to produce two distinct SMRTbell libraries: one containing fragments less than 6000 bp and another with fragments greater than 6000 bp. In the final step, the sequencing primer was annealed to the adapters and sequencing polymerase bound to SMRTbell libraries. Each sample was sequenced on a single SMRT cell. The subreads generated by the SMRT analysis tools v2.3.0 were aligned back to the reference genome, using the BLAT search genome browser.

#### End-to-end PCR

Based on the PacBio sequencing results, we designed primers overlapping the 5’ starts and the 3’ ends of the divRNAs, and we performed end-to-end PCR, using Platinum® Taq DNA Polymerase High Fidelity (Thermo Fisher Scientific). Nested PCR was then performed on the PCR-amplicons, to confirm the presence of the divRNAs. We then performed PacBio sequencing, as described above, on the End-to-end PCR amplicons where the nested PCR confirmed the presence of divRNAs, thus defining the full-length sequences of the divRNAs.

### Quantification of divRNAs, BDNF, and BDNF-AS expression in postmortem RNAseq data

The quantification of divRNAs in postmortem data poses technical challenges, because their expression, although variable across individuals, is low in basal conditions (mean expression of divRNA in the *BDNF* locus: 0.03348 RPKM, SD: 0.0557). For this reason, we estimated expression of divRNAs by counting and normalizing the reads that are aligned to the most expressed common exon of the divRNAs of the *BDNF* locus (hg38: chr11:27722351-27722486). Such exon does not overlap any other gene in this locus, as shown by our RNAseq data and RACE-PCR. Consistently, we estimated the expression of total *BDNF* by calculating and normalizing the reads overlapping the coding exon of *BDNF* (chr11:27654893-27658585), without overlapping the *BDNF-AS* exon in this region (chr11:27658241-27658462). We measured expression of the total *BDNF-AS* gene by calculating and normalizing the reads overlapping the exon-exon junction common to all the *BDNF-AS* transcripts (chr11:27658463-27659170), without being part of the readthrough transcript that connects the divergent RNA and *BDNF-AS*. Finally, we assessed the expression of the *BDNF-AS-DT* readthrough transcript by counting and normalizing the reads that overlap the most expressed unique junction that characterizes it (hg38: chr11: 27640006-27659170).

### Mouse primary cortical neuronal cell cultures

E16.5 embryos were isolated from CD timed-pregnant females, and cortices were dissected from individual embryos in base medium. Dissociation of cortices was performed as previously described (*69*). Briefly, cortices were washed with sterile PBS and incubated for 40 min at 37°C in dissociation medium containing 10 U/ml of papain (Worthington). After quenching the enzymatic reaction with trypsin inhibitor (Sigma) and BSA, cortices were gently triturated in serum-free medium containing glutamate (Invitrogen). Neurons were plated on dishes coated with poly-L-lysine (Sigma) and laminin (Invitrogen) and cultured in Neurobasal medium containing B27 supplement (Invitrogen) and antibiotics.

### ReNCell VM cultures

The ReNcell VM neural progenitor cell (NPC) line (Millipore Sigma), derived from the ventral mesencephalon by overexpression of the myc family transcription factors in human cells, has been used for stem cell therapy and disease modeling (*47, 70*). The ReNcell VM line was maintained in ReNCell NSC Maintenance medium (Millipore Sigma), supplemented with Antibiotic-Antimycotic (Gibco), 20 ng/ml EGF (Peprotech) and 20 ng/ml bFGF (Peprotech).

Cells were cultivated on 6, 12, or 24-well plates, or T75 culture flasks (Fisher Scientific), or CytoView MEA plates (Axion) pre-coated with 20 µg/ml laminin (Sigma-Aldrich). Maintenance medium was exchanged every other day until 80% cell confluence was reached, and standard differentiation was performed by removing EGF and bFGF from culture medium. After differentiation, half-medium changes were performed every 2-3 days for two weeks with maintenance medium lacking growth factors. KCl time courses were performed after complete differentiation at Day *In Vitro* (DIV) 15. Transduction with CRISPR constructs was performed at the same time as differentiation.

### Immunocytochemistry

For staining cultured ReNcell VM NPC line, samples were fixed with ice-cold 4% paraformaldehyde for 10 min. Permeabilization and blocking were performed simultaneously with 5% normal goat serum. Primary antibodies directed against FLAG (dCas-VPR), mCherry (sgRNA), and MAP2 (Millipore Sigma) were prepared in blocking buffer and incubated with the sample at 4 °C overnight. Fluorescently conjugated secondary antibodies were prepared in blocking buffer and incubated with the sample at room temperature for 90 min. Samples were counterstained with DAPI and stored in PBS until imaging.

### Neuronal depolarization experiment

To analyze whether the expression of the divRNAs in the *BDNF* locus is affected by neuronal activity in vitro, we performed a KCl-induced depolarization experiment using, in human, immortalized human neural progenitor cell line from the ventral mesencephalon (ReNcell VM NPC (*46*) and, in mouse, cortical neurons obtained from E14 mouse embryo cortices.

In human ReNcell VM NPC (*46*)), we induced neuronal depolarization by incubation in 100 mM KCl for 0 hours (h), 6 h, 6h + 3h washout (WO), 6h + 6h washout (WO), 6h + 12h washout (WO), 6h + 24h washout (WO). At each time point, we obtained cells for gene expression analysis. We then performed RNA extraction with RNeasy Mini Kit (Qiagen), reverse transcription to cDNA, using SuperScript First-Strand Synthesis System for RT-PCR (Invitrogen), and quantitative PCR (qPCR), using primers for *BDNF-DT*, the *BDNF-AS-DT* readthrough transcript, *BDNF* coding transcripts, and GAPDH as housekeeping gene.

In mouse, E14 embryo cortices were dissected and then dissociated in HBSS (Lifetech), papain and L-cysteine, using a papain dissociation system (Worthington), according to the manufacturer’s instructions. Neurons were seeded at an appropriate density for further experiments on poly-ornithine/fibronectin coated plates (VWR), in Neurobasal media (LifeTech) containing 5% Fetal Bovine Serum (LifeTech), 1% Glutamax and 1X B27 supplements (LifeTech), 1X Penicillin-Streptomycin (LifeTech). Neurons were then grown in vitro for 7 more days in the same media but without Fetal Bovine Serum. We then induced neuron depolarization, by incubation in 100 mM KCl for 0h, 3h, 6h, 6h + 3h washout (WO), 6h + 6h WO, 6h + 12h WO, 6h + 24h WO, 6h + 48h WO and, at each time point, we obtained cells for gene expression analysis. We then performed RNA extraction with RNeasy Mini Kit (Qiagen), reverse transcription to cDNA, using SuperScript First-Strand Synthesis System for RT-PCR (Invitrogen), and quantitative PCR (qPCR), using primers for *Bdnf-DT* locus, *Bdnf* protein-coding transcripts, and GAPDH as housekeeping gene.

### CRISPR/dCas9 and sgRNA construct design

A lentivirus compatible backbone (a gift from Feng Zhang, Addgene #52961) was used to insert a dCas9-VPR (VP64-p65-Rta) cassette driven by the human synapsin 1 promoter (SYN1). SP-dCas9-VPR was a gift from George Church (Addgene #63798). A guide RNA scaffold (a gift from Charles Gersbach, Addgene #47108) was inserted into a lentivirus compatible backbone containing EF1α-mCherry. A BbsI cut site within the mCherry construct was mutated with a site-directed mutagenesis kit (NEB). Bdnf-specific sgRNA targets were reused from our previous study (*45*) or were designed using CHOPCHOP (http://chopchop.cbu.uib.no/). All CRISPR RNA (crRNA) sequences were analyzed with National Center for Biotechnology Information’s (NCBI) Basic Local Alignment Search Tool (BLAST) to ensure specificity. An extensive validation of minimal sgRNA off-target effects was conducted in our previous study (*45*). The bacterial lacZ gene was used as a non-targeting sgRNA control.

### CRISPR activation of BDNF transcripts in ReNcell VM culture

Here we describe a novel iteration of the ReNcell VM line with constitutively expressed CRISPR activation (CRISPRa) system for convenient upregulation of desired target genes. ReNcell VM cultures were transduced with high titer lentivirus (multiplicity of infection (MOI) over 10,000) carrying deactivated Cas9 (dCas9) endonuclease fused to a strong transcriptional activator, VPR (VP64-p65-Rta). Successfully transduced cells were sub-cultured and passaged to create a stably expressing VPR-ReNcell VM line (**Fig. S6**). As the traditional ReNcell VM cultures, VPR-ReNcell VM cultures were differentiated into neurons by the removal of growth factors (EGF and bFGF) from the culture media for 15 DIV. Stably-transduced VPR-ReNCell VM cultures were propagated and transduced again with single guide RNA (sgRNA) lentivirus (MOI over 5,000) and differentiated into neurons.

Following CRISPRa of *BDNF*I and IV promoters, we performed RNA extraction with RNeasy Mini Kit (Qiagen), reverse transcription to cDNA, using SuperScript First-Strand Synthesis System for RT-PCR (Invitrogen), and quantitative PCR (qPCR), using primers for *BDNF-DT*, the *BDNF-AS-DT* readthrough transcript, *BDNF* coding transcripts, and GAPDH as housekeeping gene (**Fig. S4**).

### Statistical analyses

All the statistical analyses were performed in the R computing environment (*71*), version 4.3.3.

- <u>Relationship between age and *BDNF-DT*, *BDNF-AS*, *BDNF-AS-DT*, *BDNF*</u>: because gene expression does not change linearly with age, we created a spline function to modeling the relationship between expression and age, establishing knots at ages associated with evident gene expression changes, that is, 1 year, 12 years, 25 years. We assessed the significance of the relationship with age by calculating an F-statistics and a p-value from the comparison of a full spline model with age and covariates as predictors, and a covariates-only model. Covariates included in this analysis were: ancestry genetic Principal Components (PCs), and sex, as in previous work (*29*).
- <u>Relationship between *BDNF-DT*, *BDNF-AS*, *BDNF-AS-DT*, *BDNF* expression:</u> we used a linear model to analyze how the expression of these genes were each other related, adjusting for sex, age, ancestry PCs, and surrogate variables (SVs) accounting for RNA quality (*29, 34, 72*), in the sample of adult neurotypical subjects (age>13).
- <u>Differential expression analysis</u> in patients with schizophrenia and neurotypical controls was performed, in the sample with age >13, using a linear model with expression as dependent variable, and case-control status as predictors, with age, sex, ancestry PCs and SVs accounting for RNA quality (*29, 34, 72*) as covariates.
- <u>Summary-based Mendelian Randomization (SMR):</u> we performed SMR analysis(*51*) focusing on the gene expression features in the *BDNF* locus (chr11: 27,403,033-27,759,030). First, we conducted SMR using rs6265 as an instrumental variable. Next, we performed SMR without pre-selecting a candidate SNP. For each feature, we clumped cis-eQTL for each feature (including exons and splice junctions) using PLINK (500 kb window, r^2^ > 0.1). We selected independent eQTL clusters including the top eQTL (nominal p<0.05) and the top PGC3 GWAS SNPs (p< 0.05) for SMR analysis. For each cluster, we implemented SMR and HEIDI methods, using the SMR software tool (version 1.3.1) to examine pleiotropic associations of genetic variation with feature expression in the DLPFC RNAseq dataset and case–control status in the PGC3 GWAS schizophrenia data (*49*). We performed the analyses using: i) GWAS summary statistics from the primary meta-analysis for autosomal SNPs and eQTL in the full DLPFC sample; ii) GWAS summary statistics from the meta-analysis of European ancestry cohorts for autosomal SNPs and eQTL in the DLPFC sample of European ancestry.
- <u>eQTL analysis:</u> the relationship between expression and genotypes was analyzed using a linear model with expression as dependent variable, and genotype, case-control status, age, sex, genetic PCs and surrogate variable (SVs) (*29, 34, 72*) accounting for RNA quality as covariates; we repeated the same analysis using gene expression PCs instead of SVs with analogous results. In addition, we performed this analysis stratifying by diagnosis. The R package ‘MatrixeQTL’(*73*) version 2.3 was used for this analysis.
- <u>DNA methylation analyses:</u> we analyzed whether expression of *BDNF-AS-DT* is associated with methylation of the CpG in the position chr11:27658369. For this analysis, we extracted % CpG methylation levels from a published DLPFC whole-genome bisulfite sequencing dataset (*74*) partially overlapping the DLPFC RNAseq dataset of this study (N=141, **Table S1**). The relationship between expression and methylation was analyzed using a linear model with expression as dependent variable, and methylation, case-control status, age, sex, genetic PCs, methylation PCs, and surrogate variable (SVs) accounting for RNA quality as covariates. The same model was applied to other CpGs in the *BDNF* locus. The relationship between methylation at chr11:27658369 and rs6265 genotype was analyzed using a linear model with methylation as dependent variable, and genotype, case-control status, age, sex, genetic PCs, and methylation PCs as covariates.
- <u>Co-expression gene networks associated with *BDNF-DT*, with *BDNF*, *BDNF-AS*, and *BDNF-AS-DT*, and detection of concordant and discordant pattern of associations:</u> we first performed brain coexpression network analyses using each of these genes as seed gene, in RNAseq data from DLPFC of neurotypical controls (N=318), to detect genes coexpressed with *BDNF-DT*, *BDNF, BDNF-AS*, and *BDNF-AS-DT*. Specifically, after removing low-expressed genes (mean reads per kilobase million [RPKM] <0.17) using the *expression_cutoff* function in the R package *‘jaffelab’* (version 0.99.34), we performed *voom* normalization in the *Limma* package (*75*) (release 3.19), and we then used the *lmTest* and *ebayes* functions in the package to fit the statistical models to estimate log2 fold changes, moderated t-statistics, and corresponding p values. Multiple-testing correction via the false discovery rate (FDR) was applied using the set of expressed genes (23,346 genes), to define the four subsets of genes significantly co-expressed (p*_FDR-corrected_*<0.05) respectively with *BDNF-DT*, *BDNF, BDNF-AS*, and *BDNF-AS-DT.* We investigated whether these four sets of genes are enriched for particular biological features, using QIAGEN’s Ingenuity Pathway Analysis (IPA, QIAGEN, Redwood City, CA, USA; http://www.qiagen.com/ingenuity). The software determines the pathways, biological processes, and upstream regulators, enriched for a given set of genes by considering the number of focus genes that participate in each process and the total number of genes that are known to be associated with that process in the selected reference set. We performed the Ingenuity Pathway Analysis core analysis, using default parameters (reference set: Ingenuity Knowledge Base; relationships: direct and indirect; node types: all; data sources: all; confidence: experimentally observed and high; species: human, mouse, and rat; tissues and cell lines: all; mutations: all). We chose a P value calculation based on the Benjamini– Hochberg method of accounting for multiple testing in the canonical pathway and functional analyses. We finally applied, on the moderated t-statistics from the linear models described above, a threshold-free algorithm, RRHO2 (*60*), based on a rank-rank hypergeometric overlap approach, to detect pattern of concordant and discordant association of genes coexpressed with *BDNF-DT* with genes coexpressed with *BDNF*, *BDNF-AS*, and *BDNF-AS-DT*.

## Acknowledgments

We are grateful to the Lieber and Maltz families for their visionary support that funded the analytic work of this project. We are grateful to Shizhong Han, Alexander Straub, Andrew Jaffe, Yankai Jia, Bin Xie, Yuan Gao, for their contribution to data generation. We thank the Psychiatric Genomics Consortium for providing the GWAS summary statistics, and the contributors of the Genotype-Tissue Expression (GTEx) Project for generating part of the transcriptomic data that were analyzed in this work.

## Funding

This work was supported by funding from the Lieber Institute for Brain Development. The Genotype-Tissue Expression (GTEx) Project was supported by the Common Fund of the Office of the Director of the National Institutes of Health, and by NCI, NHGRI, NHLBI, NIDA, NIMH, and NINDS. GPe and LCT were supported by R01MH123567.

## Author contributions

Conceptualization: SVB, GPu, DRW, KM, GU

Data generation: SVB, NES, JHS, GPe, LCT, KRM, SCP, JEK, TMH, KM, GU

Data analysis: SVB, GPu, SM, QC, KRM, GU

Supervision: GU

Writing—original draft: SVB, GPu, KM, GU

Writing—review & editing: all authors

## Competing interests

Authors declare that they have no competing interests

## Data and materials availability

All the data analyzed in this manuscript are publicly available and can be obtained via authorized access from the dbGaP database under accession code phs000979.v3.p2.

The GTEX data used to generate data of Supplementary Figure S3i were obtained from the GTEx Portal on 04/03/2025.

## Notes

### Competing Interest Statement

The authors have declared no competing interest.

### Author Declarations

The study used openly available postmortem human data available on the dbGaP database under accession code phs000979.v3.p2 Postmortem human brain tissue was obtained by autopsy primarily from the Offices of the Chief Medical Examiner of the District of Columbia and of the Commonwealth of Virginia, Northern District, all with informed consent from the legal next of kin (protocol 90-M-0142 approved by the NIMH/NIH Institutional Review Board). The National Institute of Child Health and Human Development Brain and Tissue Bank for Developmental Disorders (http://www.BTBank.org) provided additional postmortem prenatal, infant, child and adolescent brain tissue samples, by under contracts NO1-HD-4-3368 and NO1-HD-4-3383. The Institutional Review Board of the University of Maryland at Baltimore and the State of Maryland approved the protocol, and the tissue was donated to the Lieber Institute for Brain Development under the terms of a Material Transfer Agreement.

